# Radiographically Successful Periacetabular Osteotomy Does Not Achieve Optimal Contact Mechanics in Dysplastic Hips

**DOI:** 10.1101/2022.05.26.22275634

**Authors:** Holly D. Aitken, Aspen Miller, Dominic J.L. Rivas, Marcus Tatum, Robert W. Westermann, Michael C. Willey, Jessica E. Goetz

## Abstract

**Introduction:** Optimal correction of hip dysplasia deformity with periacetabular osteotomy (PAO) that minimizes elevated contact stresses may reduce osteoarthritis (OA) development.

**Questions/Purposes:** We used a computational approach based on discrete element analysis (DEA) to determine (1) if computational optimization can identify patient-specific acetabular corrections that optimize joint contact mechanics, (2) whether a strictly mechanically optimal correction is clinically feasible, and (3) whether the contact mechanics of optimal corrections differ from those of surgically achieved corrections.

**Methods:** Preoperative and postoperative hip models were created from CT scans of a retrospective cohort (n=20) who underwent PAO to treat hip dysplasia. A digitally extracted acetabular fragment was computationally rotated in two-degree increments of lateral and anterior coverage to simulate candidate PAO reorientations. DEA-computed contact stress for each candidate reorientation model was used to select a purely mechanically optimal reorientation that minimized chronic contact stress exposures above damaging thresholds and a clinically optimal reorientation that balanced reducing chronic exposures with achieving clinically realistic acetabular orientations. Radiographic coverage, contact area, peak/mean contact stress, and peak/mean cumulative exposure were compared between preoperative, mechanically optimal, clinically optimal, and surgically achieved acetabular orientations.

**Results:** Computationally optimal reorientations had significantly (*p*<0.001) more lateral and anterior coverage than surgically achieved PAO corrections. The mechanically/clinically optimal reorientations also had significantly more contact area (*p*<0.001*/p*=0.001) and significantly lower peak contact stress (*p*<0.001/*p*<0.001), mean contact stress (*p*<0.001*/p*=0.001), peak chronic exposure (*p*=0.001*/p*=0.003), and mean chronic exposure (*p*<0.001*/p*=0.001) than the surgically achieved corrections.

**Conclusions:** This computational approach identified patient-specific mechanically optimal and clinically optimal acetabular reorientations. Surgically achieved reorientations did not reduce contact stress exposure to the extent achieved with computed optimal reorientations. However, optimal orientations identified for many patients risk secondary femoroacetabular impingement. Identifying patient-specific corrections that balance optimizing mechanics with clinical reality is necessary to reduce the risk of OA progression after PAO.

## INTRODUCTION

Hip dysplasia, a complex array of acetabular and femoral deformities, results in joint instability and altered intra-articular mechanics [13, 26]. These pathologic mechanics frequently cause hip pain and premature development of osteoarthritis (OA) in active young adult patients [10, 25]. Adult hip dysplasia is commonly treated with a periacetabular osteotomy (PAO), a well-described surgical procedure that permits multiplanar reorientation of the acetabulum to improve deficiencies in femoral head coverage, medialize the hip joint center, and reduce elevated joint contact stress [11, 27, 35]. Reorientation of the dysplastic acetabulum via PAO can improve acetabular coverage to better resemble normal hips in terms of 2D radiographic measures [21], but joint contact stress remains elevated compared to normal hips [12]. Computational techniques based on 3D imaging may provide better assessment of optimum deformity correction with PAO.

Discrete element analysis (DEA) is a rapidly executing, highly numerically stable computational modeling technique in which articular cartilage is represented as a bed of compressive springs supported by rigid underlying bone surfaces. The amount of spring deformation that occurs under applied load is used to calculate the local tissue forces and associated contact stresses [31]. This simplified modeling technique has been shown to accurately predict contact stresses in the hip joint [1, 38]. Prior studies using DEA to investigate joint contact mechanics in dysplastic hips have demonstrated elevated contact stresses and reduced contact areas in dysplastic joints compared to normal hips [12, 25, 26] and shown that contact stress elevations are not reduced to levels of radiographically normal hips after PAO [12]. These findings suggest that radiographic normalization of the joint may be insufficient for achieving a non-pathologic mechanical environment.

There has been an increased focus on identifying patient-specific, mechanically driven, optimal acetabular corrections to improve postoperative contact mechanics [6, 7, 17, 20, 23, 39, 42, 43]. However, most of those studies [6, 7, 17, 39] discern the optimal acetabular reorientation based on peak joint contact stress, which has been shown to correlate poorly with patient-reported outcomes [6, 7, 13]. In contrast, several previous studies across numerous joints have established a correlation between osteoarthritis (OA) progression and cumulative contact stress exposure, a metric that evaluates exposure to damaging contact stresses over time [5, 13, 19, 33, 34]. Identifying patient-specific acetabular corrections that minimize a mechanical parameter associated with OA development, such as contact stress exposure, may be necessary to return the mechanical joint environment to a non-destructive state.

The purpose of this study was to use a computational optimization approach to retrospectively perform virtual PAO and DEA analysis to determine (1) whether a computational optimization approach can identify patient-specific acetabular corrections that minimize an individual’s negative contact mechanics, (2) whether a strictly mechanically optimal correction is clinically feasible, and (3) whether acetabular coverage measures and computed contact mechanics of optimal corrections are different from those of surgically achieved corrections.

## METHODS

With Institutional Review Board approval, two consecutive series of patients were retrospectively identified for this study. The first series of 10 patients underwent PAO to correct hip dysplasia by a single surgeon (MCW) between January 2018 and September 2018. The second consecutive series of 10 patients underwent PAO (MCW) to correct acetabular dysplasia with concurrent hip arthroscopy for correction of femoral head-neck offset deformity by a single surgeon (RWW) between January 2019 and May 2019. Patients missing preoperative or postoperative CT or radiographic imaging and those with severe joint incongruency that prevented cartilage projection with current modeling techniques were excluded from these otherwise consecutive series.

### Model Generation

To create patient-specific hip models, the pelvic, proximal femoral, and distal femoral bony geometry was segmented from preoperative and postoperative CT scans using MIMICS image processing software (Materialise, Plymouth, MI). Using a previously described methodology [2], articular cartilage was approximated in these models by projecting the acetabular and femoral subchondral bone surfaces a distance corresponding to the patient’s joint space and custom smoothing these projected surfaces. Each model was oriented to a standardized hip joint coordinate system defined using bony anatomic landmarks [41] and loaded using the average hip joint reaction forces and rotation angles derived from gait analysis and musculoskeletal modeling of walking gait in a series of 10 patients with hip dysplasia [15]. These average joint reaction forces and hip rotation angles were discretized into 13 quasi-static time points spanning the stance phase of walking gait and scaled based on the individual patient’s bodyweight for application to each patient’s model. Cartilage was assigned isotropic, linear-elastic material properties (E = 8 MPa, ν = 0.42) [2, 38], and labral/capsular restraint was modeled as a linear spring resisting medial-lateral translation of the pelvis (spring constant = 55 N/mm) [2].

### Chronic Contact Stress Exposure

DEA calculations were performed using a previously developed Newton’s method solver implemented in MATLAB [19]. To identify acetabular corrections that minimize chronic joint loading above levels associated with OA development, a 1 MPa damage threshold previously shown to be correlated with intra-articular cartilage degeneration [2] was used to isolate contact stresses considered damaging to the joint. These damaging contact stresses were then multiplied by the length of time spent in each loading configuration and summed across the 13 configurations comprising the gait cycle [5]. Each per-step contact stress-time summation was then scaled by an assumed 2 million walking steps per year [16, 30] and multiplied by the patient’s age in years to obtain the chronic contact stress-time exposure for that patient [2]. Chronic contact stress-time exposures over a 2 MPa-years accumulated damage threshold that has been associated with cartilage degeneration [2] were considered detrimental to the health of the joint.

### Computational Optimization

Virtual PAO was simulated for each patient by manually isolating the acetabular region from the preoperative pelvis surface using a series of planes to approximate the intraoperative PAO osteotomies (Figure 1). A custom algorithm developed in MATLAB (Mathworks, Natick, MA) computationally rotated the extracted acetabular fragment into candidate PAO reorientations. Lateral and anterior coverage were modified by rotating the acetabular fragment around rotational axes specified based on anatomic bony landmarks and located at the center of a best-fit sphere to the acetabular subchondral bone surface (Figure 1).

**Figure 1:**
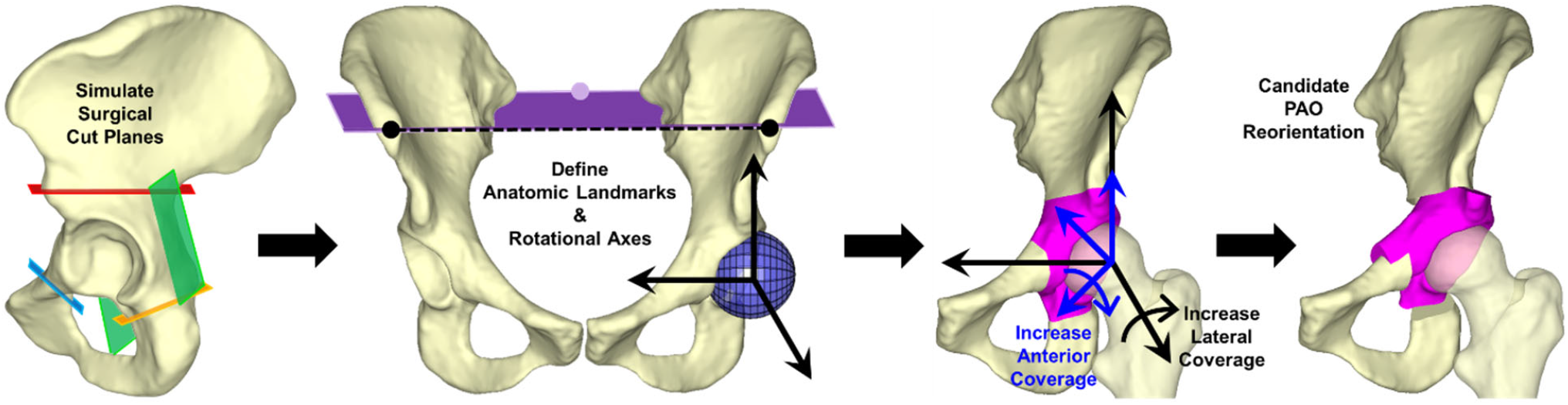
(Left) From each patient’s preoperative hip model, a simulated acetabular fragment was extracted from the remainder of the preoperative pelvis surface geometry using a series of planes approximating the intraoperative PAO osteotomies. (Left center) Lateral coverage changes were simulated by rotating the extracted fragment about an anteroposterior axis orthogonal to both (1) a line (dashed black) defined between the left and right anterior superior iliac spines (ASISs) (black dots) and (2) a line perpendicular to the plane (purple) defined by the left and right ASISs and the midpoint between the left and right posterior superior iliac spines (purple dot). This anteroposterior axis was located at the center of a best-fit sphere to the acetabular subchondral bone (blue sphere). (Right center) Anterior coverage of the acetabulum was increased by rotating the fragment around an oblique axis (blue coordinate system) defined by rotating the anteroposterior axis defined for lateral coverage (black coordinate system) 45 degrees in the transverse plane about the vertical axis (clockwise rotation for left acetabula, counterclockwise rotation for right acetabula). (Right) Varying levels of lateral and anterior coverage were simulated as candidate PAO reorientations.

Candidate reorientations were generated in two-degree increments through clinically acceptable combinations of normal lateral (24-44 degrees) and anterior (20-44 degrees) coverage [9, 28] (Figure 2). The reoriented acetabular fragment was combined with the preoperative femur surface to create each candidate PAO model. For each of the total 143 candidate reorientations, DEA was used to compute contact stresses and chronic contact stress-time exposures.

**Figure 2:**
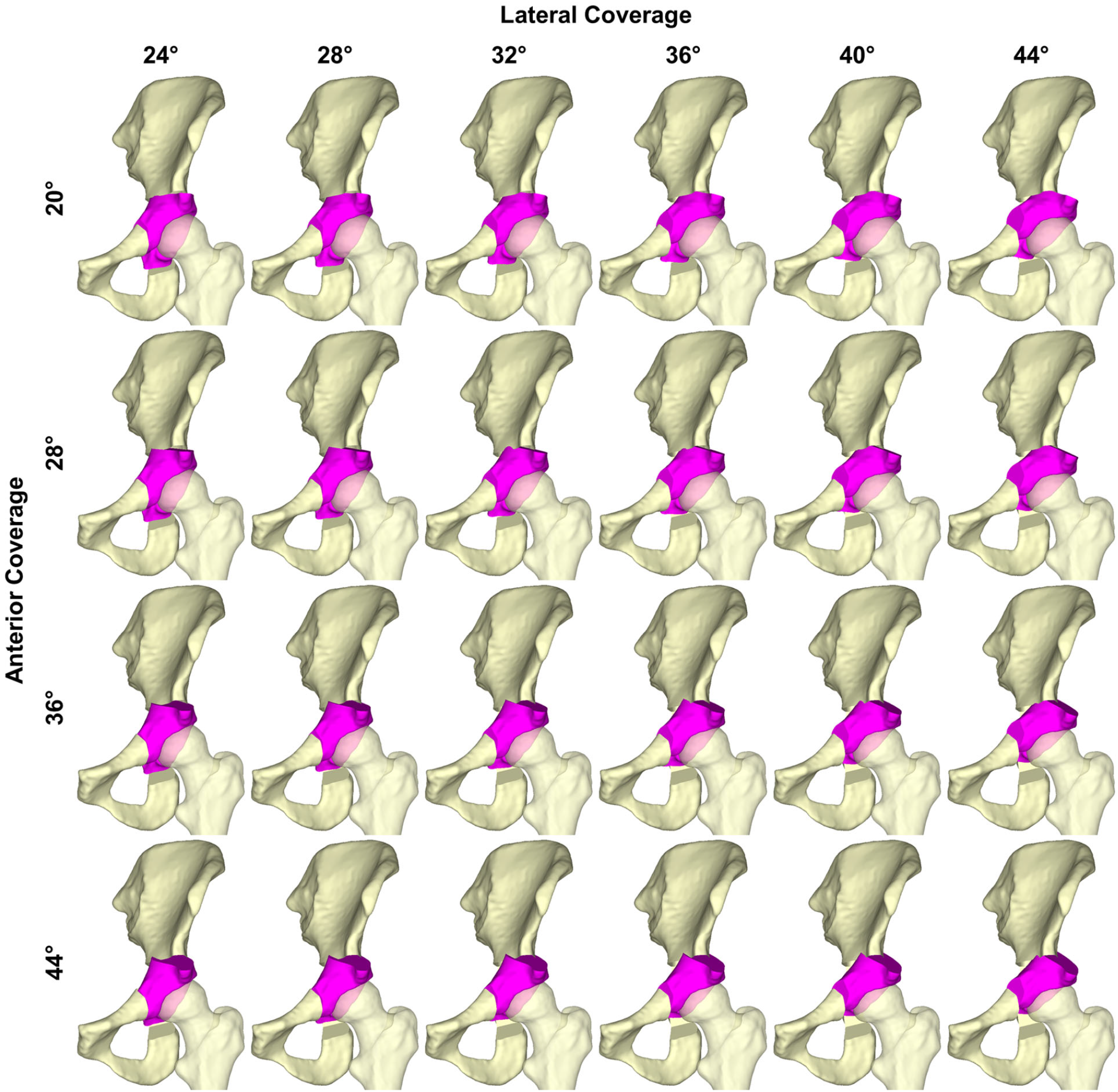
AP view of candidate PAO reorientations investigated by computational optimization for a representative dysplastic hip model. The algorithm rotates the acetabular fragment (pink) in two-degree increments of both lateral and anterior coverage, resulting in a total of 143 candidate reorientations being investigated. To better illustrate the differences between acetabular fragment rotations, only every 4 degrees of lateral coverage and 8 degrees of anterior coverage are shown.

To identify the optimal PAO reorientation from the candidates, a cost function, referred to as the mechanically optimal exposure (Appendix), was developed that incorporated the magnitude, surface area, and regional location of chronic contact stress-time exposures over the 2 MPa-years accumulated damage threshold [2]. Weights were assigned to each of six acetabular regions to prioritize shifting contact towards the medial acetabulum and reduce damaging chronic over-exposures along the lateral acetabular rim. While the mechanically optimal exposure metric provides a cost function that can be numerically minimized, an optimization strategy based solely on minimizing these mechanical criteria may select acetabular reorientations that would cause severe impingement with limited hip range of motion and are thus unrealistic for clinical implementation. To ensure that the optimization approach was identifying mechanically optimized and more surgically acceptable reorientations, a second cost function, referred to as the clinically optimal exposure (Appendix), was developed that balanced optimizing chronic mechanical exposures with clinical reality through the addition of a surgically allowable rotational term. Optimal reorientations were selected using both the mechanically optimal exposure *fmechanical* and the clinically optimal exposure *f*_*clinical*_.

### Radiographic Coverage Assessment

When comparing acetabular coverage of the computationally optimal reorientations to surgically achieved coverage, a 1-degree change in radiographic coverage measurements (LCEA, ACEA) did not directly equate to a 1-degree rotation around the specific rotational axes defined for optimization. Therefore, radiographic coverage of the optimal reorientations was measured on digitally reconstructed radiographs (DRRs) of each patient’s preoperative hip model with the simulated acetabular fragment in the optimized position (Figure 3). To create the DRRs, a custom MATLAB algorithm [18] generated a virtual “radiograph projector” that created 2D projections of the 3D hip model that matched the viewing angle, resolution, detector size, and source-to-detector distance of the patient’s preoperative clinical radiographs. Then, the viewing vector through the 3D model was manually adjusted until the bony geometry in the 2D projections matched the bony geometry in the patient’s preoperative clinical radiographs. Once the pelvis was aligned, the simulated acetabular fragment was reoriented to the mechanically and clinically optimal reorientations relative to the pelvis. Visually accurate DRRs were created by converting the Hounsfield units from the CT scan used to generate the 3D model into linear attenuation coefficients to accurately recreate fluoroscopic image intensity values [8]. To view overlapping bones in the same DRR, the algorithm used the concept of multi-bone DRR generation, in which the intensity values of the bone of interest (here the acetabular fragment) and neighboring bones (here the femur and remainder of the hemipelvis) contribute to the intensity values where they overlap in the DRR [14]. Lateral acetabular coverage (LCEA) and anterior acetabular coverage (ACEA) were measured on anteroposterior and false profile DRRs of the optimal acetabular reorientations and on the preoperative and postoperative clinical radiographs by two trained individuals (MT, MCW). The average of the two individuals’ measurements was considered the final radiographic measurement.

**Figure 3:**
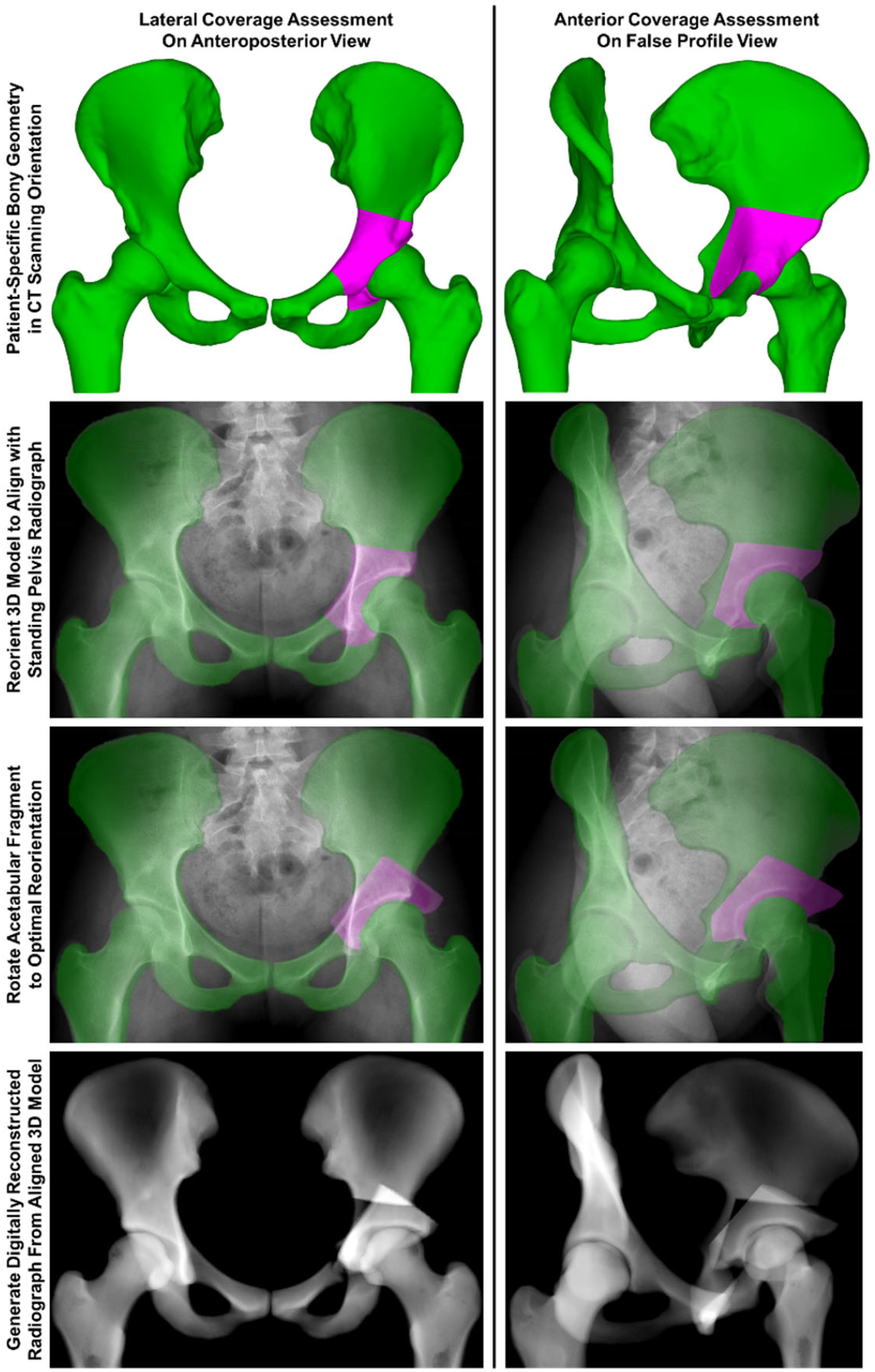
Process of generating digitally reconstructed radiographs (DRRs) from each patient-specific model for assessing lateral (left column) and anterior (right column) acetabular coverage. The patient-specific model geometry (top row) is reoriented to align with the pelvis orientation in the patient’s preoperative standing AP and false profile pelvis radiographs (second row). The simulated acetabular fragment (pink) is then rotated to the computationally identified optimal acetabular orientation (third row). The aligned patient-specific geometry (green) with the acetabular fragment in the optimal orientation (pink) is then used to generate the DRRs (bottom row) used to assess LCEA and ACEA, traditional radiographic measures of acetabular coverage.

### Statistical Analysis

Radiographic coverage, contact stress, chronic contact stress exposure, and contact area were compared between preoperative, mechanically optimal, clinically optimal, and surgically achieved acetabular orientations using Wilcoxon matched-pairs signed-rank tests. Data are presented as medians [interquartile ranges (IQR)], and statistical significance was set at *p* ≤ 0.05 with a Holm-Bonferroni correction for multiple comparisons. Intraclass correlation coefficients (ICCs) were used to describe interrater reliability in radiographic measurements. All statistical analyses were completed in SAS 9.4 (SAS Institute Inc, Cary, NC, USA).

## RESULTS

Both the mechanically optimal and clinically optimal exposure metrics were able to identify unique, individualized acetabular orientations that minimized negative contact mechanics for each patient’s hip model (Table 1). Compared to the preoperative condition, mechanically/clinically optimal orientations predicted significantly lower peak contact stress (*p* < 0.001/*p* = 0.001), mean contact stress (*p* < 0.001/*p* = 0.001), peak chronic exposure (*p* = 0.001/*p* = 0.005), mean chronic exposure (*p* < 0.001/*p* < 0.001), and higher contact area (*p* < 0.001/*p* < 0.001). As expected, compared to the clinically optimal acetabular orientation, the mechanically optimal acetabular orientations had significantly lower peak/mean contact stress (*p* = 0.004/*p* = 0.001), lower peak/mean chronic exposure (*p* = 0.003/*p* = 0.001), and higher contact area (*p* = 0.002).

**Table 1:**
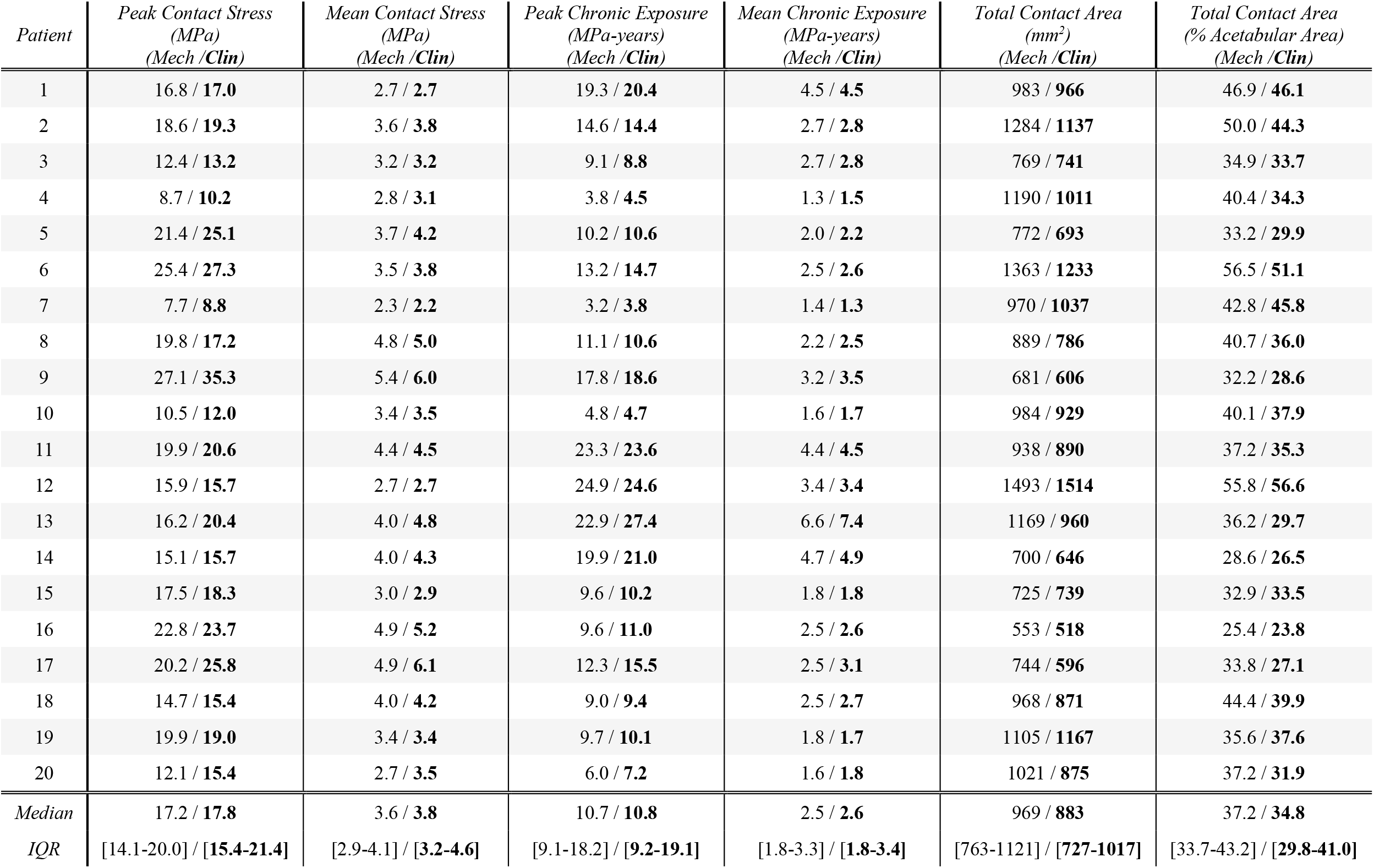
Optimal contact mechanics were unique for each patient model when utilizing both the mechanically optimal (Mech) and clinically optimal (Clin) acetabular orientations.

| *Patient* | *Peak Contact Stress*<br>*(MPa)*<br>*(Mech /Clin)* | *Mean Contact Stress*<br>*(MPa)*<br>*(Mech /Clin)* | *Peak Chronic Exposure*<br>*(MPa-years)*<br>*(Mech /Clin)* | *Mean Chronic Exposure*<br>*(MPa-years)*<br>*(Mech /Clin)* | *Total Contact Area*<br>*(mm<sup>2</sup>)*<br>*(Mech /Clin)* | *Total Contact Area*<br>*(% Acetabular Area)*<br>*(Mech /Clin)* |
| --- | --- | --- | --- | --- | --- | --- |
| 1 | 16.8 / **17.0** | 2.7 / **2.7** | 19.3 / **20.4** | 4.5 / **4.5** | 983 / **966** | 46.9 / **46.1** |
| 2 | 18.6 / **19.3** | 3.6 / **3.8** | 14.6 / **14.4** | 2.7 / **2.8** | 1284 / **1137** | 50.0 / **44.3** |
| 3 | 12.4 / **13.2** | 3.2 / **3.2** | 9.1 / **8.8** | 2.7 / **2.8** | 769 / **741** | 34.9 / **33.7** |
| 4 | 8.7 / **10.2** | 2.8 / **3.1** | 3.8 / **4.5** | 1.3 / **1.5** | 1190 / **1011** | 40.4 / **34.3** |
| 5 | 21.4 / **25.1** | 3.7 / **4.2** | 10.2 / **10.6** | 2.0 / **2.2** | 772 / **693** | 33.2 / **29.9** |
| 6 | 25.4 / **27.3** | 3.5 / **3.8** | 13.2 / **14.7** | 2.5 / **2.6** | 1363 / **1233** | 56.5 / **51.1** |
| 7 | 7.7 / **8.8** | 2.3 / **2.2** | 3.2 / **3.8** | 1.4 / **1.3** | 970 / **1037** | 42.8 / **45.8** |
| 8 | 19.8 / **17.2** | 4.8 / **5.0** | 11.1 / **10.6** | 2.2 / **2.5** | 889 / **786** | 40.7 / **36.0** |
| 9 | 27.1 / **35.3** | 5.4 / **6.0** | 17.8 / **18.6** | 3.2 / **3.5** | 681 / **606** | 32.2 / **28.6** |
| 10 | 10.5 / **12.0** | 3.4 / **3.5** | 4.8 / **4.7** | 1.6 / **1.7** | 984 / **929** | 40.1 / **37.9** |
| 11 | 19.9 / **20.6** | 4.4 / **4.5** | 23.3 / **23.6** | 4.4 / **4.5** | 938 / **890** | 37.2 / **35.3** |
| 12 | 15.9 / **15.7** | 2.7 / **2.7** | 24.9 / **24.6** | 3.4 / **3.4** | 1493 / **1514** | 55.8 / **56.6** |
| 13 | 16.2 / **20.4** | 4.0 / **4.8** | 22.9 / **27.4** | 6.6 / **7.4** | 1169 / **960** | 36.2 / **29.7** |
| 14 | 15.1 / **15.7** | 4.0 / **4.3** | 19.9 / **21.0** | 4.7 / **4.9** | 700 / **646** | 28.6 / **26.5** |
| 15 | 17.5 / **18.3** | 3.0 / **2.9** | 9.6 / **10.2** | 1.8 / **1.8** | 725 / **739** | 32.9 / **33.5** |
| 16 | 22.8 / **23.7** | 4.9 / **5.2** | 9.6 / **11.0** | 2.5 / **2.6** | 553 / **518** | 25.4 / **23.8** |
| 17 | 20.2 / **25.8** | 4.9 / **6.1** | 12.3 / **15.5** | 2.5 / **3.1** | 744 / **596** | 33.8 / **27.1** |
| 18 | 14.7 / **15.4** | 4.0 / **4.2** | 9.0 / **9.4** | 2.5 / **2.7** | 968 / **871** | 44.4 / **39.9** |
| 19 | 19.9 / **19.0** | 3.4 / **3.4** | 9.7 / **10.1** | 1.8 / **1.7** | 1105 / **1167** | 35.6 / **37.6** |
| 20 | 12.1 / **15.4** | 2.7 / **3.5** | 6.0 / **7.2** | 1.6 / **1.8** | 1021 / **875** | 37.2 / **31.9** |
| *Median* | 17.2 / **17.8** | 3.6 / **3.8** | 10.7 / **10.8** | 2.5 / **2.6** | 969 / **883** | 37.2 / **34.8** |
| *IQR* | [14.1-20.0] / **[15.4-21.4]** | [2.9-4.1] / **[3.2-4.6]** | [9.1-18.2] / **[9.2-19.1]** | [1.8-3.3] / **[1.8-3.4]** | [763-1121] / **[727-1017]** | [33.7-43.2] / **[29.8-41.0]** |

Interrater reliability of lateral coverage measurement was excellent on the mechanically optimal reorientation images (ICC = 0.905, 95% CI = [0.779-0.961]) and good on the clinically optimal reorientations (ICC = 0.865, 95% CI = [0.697-0.944]). Similarly, there was good reliability in anterior coverage measurement for the mechanically optimal reorientations (ICC = 0.812, 95% CI = [0.589-0.921]) and moderate reliability for the clinically optimal reorientations (ICC = 0.671, 95% CI = [0.299-0.860]). Using the mechanically optimal exposure metric frequently predicted optimal acetabular orientations with median lateral coverage of 46 degrees [42-48 degrees] and median anterior coverage of 53 degrees [48-60 degrees], which would be considered clinically over-corrected (Table 2 and Figure 4). Incorporating the surgical rotation term into calculation of the clinically optimal exposure identified optimal acetabular reorientations that were more clinically realistic with significantly less lateral (40 degrees [37-45 degrees], *p* = 0.011) and anterior (46 degrees [42-48 degrees], *p* < 0.001) coverage compared to those predicted using the mechanically optimal exposure, but these rotations would still be considered clinically over-corrected (Table 2).

**Table 2:** Lateral and anterior coverage values measured radiographically for the preoperative, mechanically optimal, clinically optimal, and surgically achieved acetabular orientations for each patient model.

| Patient | *Lateral Acetabular Coverage (LCEA)* |  |  |  | *Anterior Acetabular Coverage (ACEA)* |  |  |  |
| --- | --- | --- | --- | --- | --- | --- | --- | --- |
|  | *Preoperative Orientation* | *Mechanically Optimal Reorientation* | *Clinically Optimal Reorientation* | *Surgically Achieved Reorientation* | *Preoperative Orientation* | *Mechanically Optimal Reorientation* | *Clinically Optimal Reorientation* | *Surgically Achieved Reorientation* |
| 1 | 11 | 47 | 50 | 31 | 15 | 65 | 56 | 35 |
| 2 | -3 | 44 | 37 | 14 | -7 | 62 | 45 | 10 |
| 3 | 19 | 51 | 47 | 31 | 23 | 56 | 47 | 37 |
| 4 | 13 | 39 | 37 | 35 | 13 | 60 | 38 | 28 |
| 5 | 21 | 43 | 40 | 29 | 19 | 62 | 50 | 35 |
| 6 | 22 | 55 | 49 | 32 | 12 | 58 | 42 | 31 |
| 7 | 19 | 32 | 37 | 34 | 15 | 40 | 42 | 34 |
| 8 | 18 | 43 | 37 | 30 | 17 | 39 | 38 | 37 |
| 9 | 20 | 46 | 38 | 30 | 36 | 51 | 44 | 35 |
| 10 | 16 | 48 | 48 | 34 | 14 | 52 | 47 | 36 |
| 11 | 25 | 49 | 46 | 39 | 24 | 47 | 46 | 48 |
| 12 | 20 | 36 | 38 | 37 | 24 | 41 | 41 | 41 |
| 13 | 16 | 52 | 45 | 35 | 14 | 70 | 51 | 29 |
| 14 | 22 | 45 | 42 | 35 | 24 | 64 | 62 | 41 |
| 15 | 17 | 33 | 35 | 35 | 27 | 48 | 44 | 43 |
| 16 | 22 | 47 | 42 | 36 | 27 | 54 | 46 | 39 |
| 17 | 33 | 46 | 40 | 42 | 55 | 53 | 46 | 43 |
| 18 | 18 | 47 | 45 | 34 | 25 | 59 | 58 | 37 |
| 19 | 15 | 34 | 39 | 33 | 18 | 41 | 45 | 38 |
| 20 | 30 | 48 | 35 | 33 | 43 | 50 | 41 | 38 |
| *Median* | 19 | 46 | 40 | 34 | 21 | 53 | 46 | 37 |
| *IQR* | [16-22] | [42-48] | [37-45] | [31-35] | [15-26] | [48-60] | [42-48] | [35-39] |

**Figure 4:**
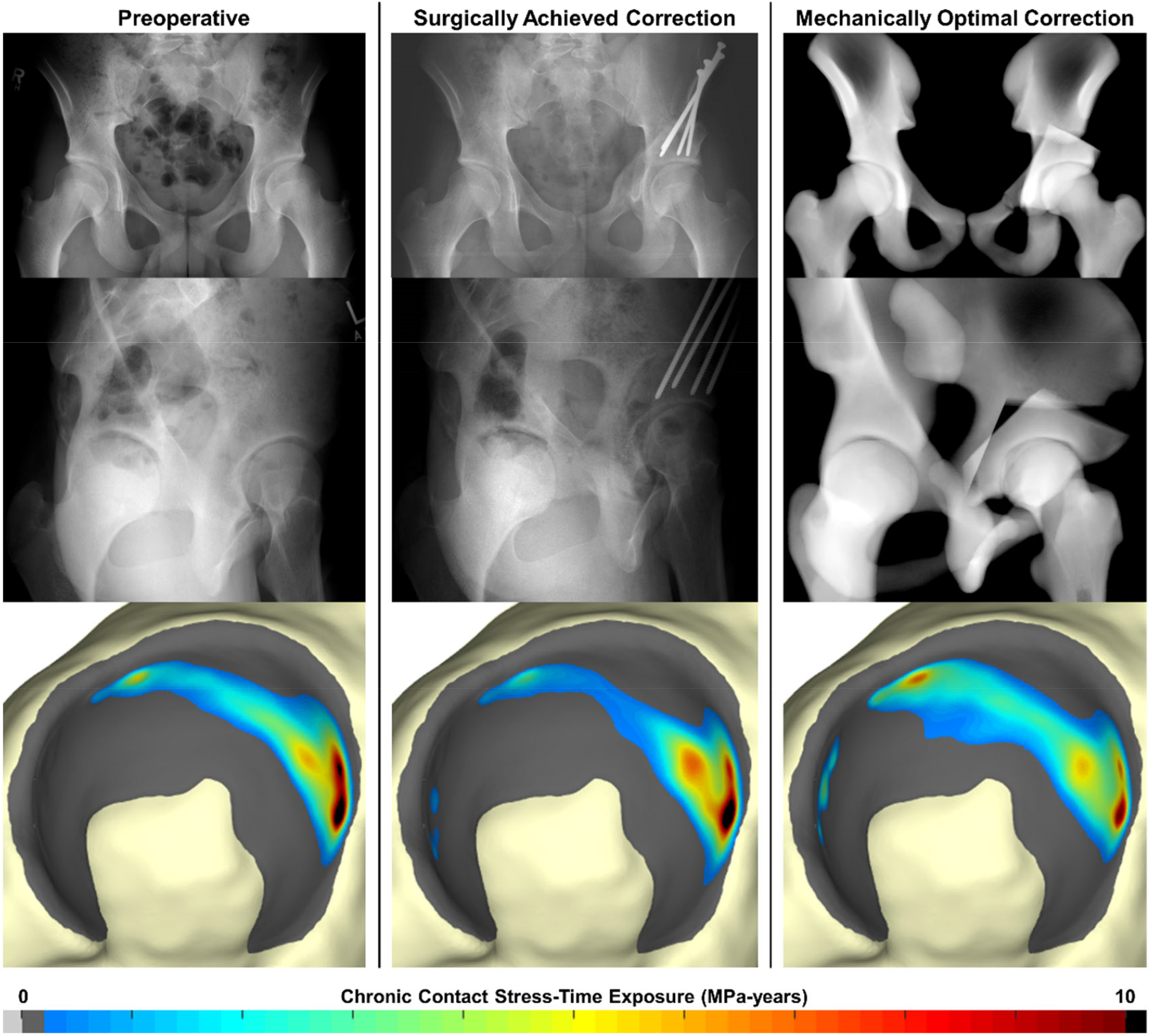
(Left column) llustrative example of a dysplastic hip with radiographically measured lateral coverage of 15 degrees and anterior coverage of 18 degrees preoperatively. Chronic contact stress-time exposure is located primarily along the lateral edge of the acetabulum. (Center column) Radiographic coverage was surgically corrected to 33 degrees of lateral coverage and 38 degrees of anterior coverage, thereby radiographically normalizing acetabular coverage but only slightly improving chronic contact stress-time exposure. (Right column) The mechanically optimal correction predicted slightly greater acetabular coverage of 34 degrees laterally and 41 degrees anteriorly, which would be clinically considered slight over-coverage, but significantly medialized the contact region of the acetabulum and reduced chronic contact stress-time exposure.

Surgically achieved PAO corrections had significantly less lateral (34 degrees [31-35 degrees]) and anterior (37 degrees [35-39 degrees]) coverage compared to both the mechanically optimal reorientations (lateral: 46 degrees [42-48 degrees], *p* < 0.001; anterior: 53 degrees [48-60 degrees], *p* < 0.001) and clinically optimal reorientations (lateral: 40 degrees [37-45 degrees], *p* < 0.001; anterior: 46 degrees [42-48 degrees], *p* < 0.001) (Table 2). Peak and mean contact stress was significantly higher in the surgically achieved PAO corrections (peak stress: 24.2 MPa [18.9-30.3 MPa]; mean stress: 5.1 MPa [4.2-6.1 MPa]) compared to the mechanically optimal (peak stress: 17.2 MPa [14.1-20.0 MPa], *p* < 0.001; mean stress: 3.6 MPa [2.9-4.1 MPa], *p* < 0.001) and the clinically optimal (peak stress: 17.8 MPa [15.4-21.4 MPa], *p* < 0.001; mean stress: 3.8 MPa [3.2-4.6 MPa], *p* = 0.001) reorientations (Figure 5, top row). Similarly, surgically achieved reorientations had significantly higher peak (14.1 MPa-years [11.6-19.6 MPa-years]) and mean (3.1 MPa-years [2.2-3.9 MPa-years]) chronic exposure than mechanically optimal (peak exposure: 10.7 MPa-years [9.1-18.2 MPa-years], *p* = 0.001; mean exposure: 2.5 MPa-years [1.8-3.3 MPa-years], *p* < 0.001) or the clinically optimal (peak exposure: 10.8 MPa-years [9.2-19.1 MPa-years], *p* = 0.003; mean exposure: 2.6 MPa-years [1.8-3.4 MPa-years], *p* = 0.001) reorientations (Figure 5, center row & Figure 6). Total contact area was significantly less in the surgically achieved PAO corrections (644 mm^2^ [576-843 mm^2^]) than in the mechanically optimal (969 mm^2^ [763-1121 mm^2^], *p* < 0.001) or the clinically optimal (883 mm^2^ [727-1017 mm^2^], *p* = 0.001) reorientations (Figure 5, bottom row).

**Figure 5:**
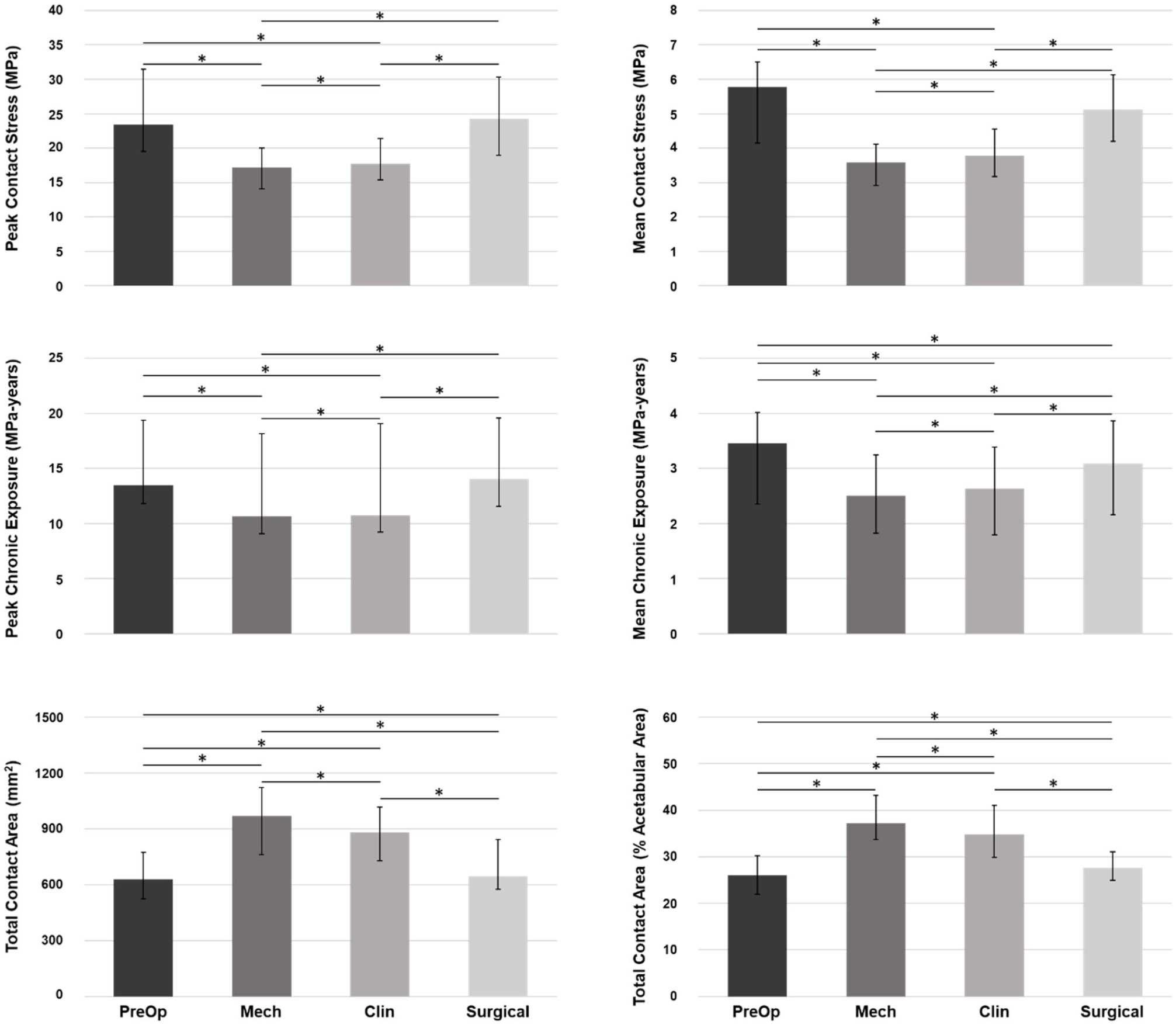
(Top left) Peak contact stress, (top right) mean contact stress, (center left) peak chronic contact stress-time exposure, (center right) mean chronic contact stress-time exposure, and (bottom left and bottom right) total contact area when the acetabular fragment is oriented in the preoperative, mechanically optimal, clinically optimal, and surgically achieved orientations. Bars indicate the median of the 20 hip models, and error bars indicate the interquartile range (IQR). * indicates *p* < 0.05 after Holm-Bonferroni correction.

**Figure 6:**
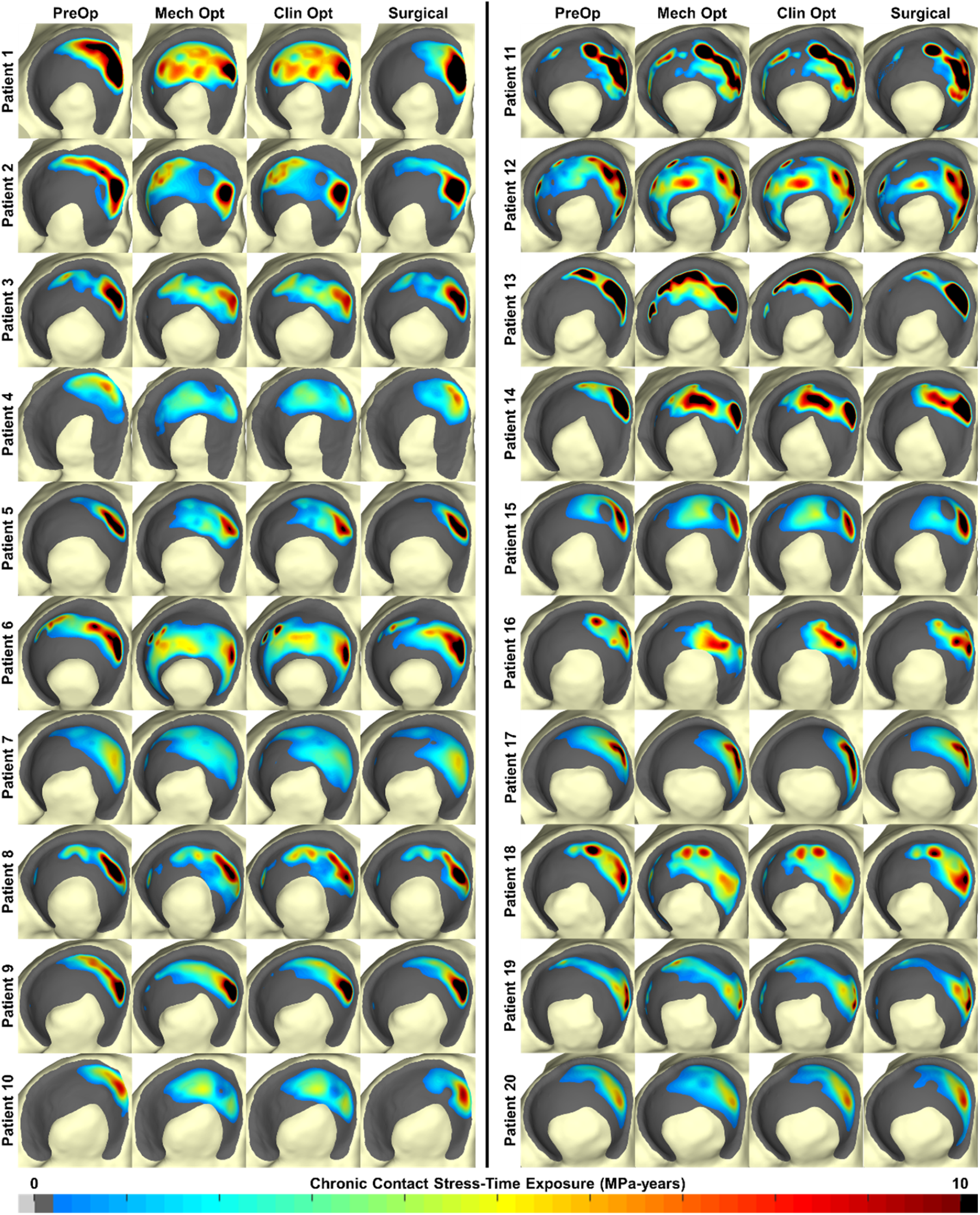
Chronic contact stress-time exposure distributions for each patient model when the acetabular fragment is in the preoperative, mechanically optimal, clinically optimal, and surgically achieved orientations. Anterior is on the right for all acetabula. The surgically achieved reorientations improve the chronic exposure distributions relative to preoperative but do not achieve optimal contact mechanics.

## DISCUSSION

The objectives of performing PAO for treatment of adult hip dysplasia are to reorient a shallow, upturned acetabulum to improve joint stability, reduce elevated joint contact stress, and prevent or reduce OA development. While PAO allows surgeons to normalize the acetabular orientation according to radiographic measures [21], the postoperative acetabular orientation may fail to improve, or even worsen, local contact stress elevations [12]. In this study, a computational optimization approach was used to retrospectively identify patient-specific acetabular orientations that would minimize detrimental joint contact mechanics. Using a cost function that simply minimized contact stress exposure frequently resulted in predicted optimal acetabular orientations that would be considered clinically over-corrected. The addition of a surgically allowable rotational term resulted in prediction of optimal orientations that were slightly more clinically realistic. Contact stress and chronic contact stress-time exposure was significantly less and total contact area was significantly higher in the clinically optimal orientations compared to surgically achieved corrections. Interestingly, some clinically optimal reorientations predicted less acetabular coverage than that achieved surgically, indicating that maximizing coverage or achieving a radiographically normal correction may not guarantee optimal contact mechanics after PAO.

Previous computational studies aiming to assess how planned acetabular reorientations alter contact mechanics in dysplastic hips have been subjected to numerous limitations that reduce their clinical applicability. The majority of those optimization studies [6, 7, 17, 39] have attempted to use peak contact stress to discern the best acetabular reorientation, even though peak contact stress has been shown to correlate poorly with patient-reported outcome measures [6, 7, 13]. To our knowledge, we are the first to identify optimal acetabular corrections using a metric of chronic joint loading above levels identified as being associated with intra-articular cartilage damage [2]. Further, the computational models used in optimization studies of dysplastic hips have implemented numerous modeling simplifications, such as assuming spherical joint geometry [17], uniform cartilage thickness [7], and non-dysplastic walking gait [23, 24, 39], that greatly affect their accuracy in the young adult hip dysplasia population. The cartilage generation technique used in this work has undergone extensive validation against pressure calculations in cadaveric specimens [2], and the kinematic and kinetic inputs used to load the DEA models were obtained from walking gait in patients with hip dysplasia [15], thereby drastically improving the applicability of the contact stress computations used to identify optimal acetabular corrections.

This relatively small cohort of 20 consecutive patients does contains some notable outliers, both in terms of the degree of preoperative deformity and the amount of coverage correction, that significantly affected the median acetabular coverage and mechanical results. For instance, one case had a preoperative LCEA of -3 degrees and ACEA of -7 degrees, indicating significant dysplastic deformity. During surgery, PAO was able to substantially increase acetabular coverage in this patient to a LCEA of 14 degrees and ACEA of 10 degrees. However, this surgically achieved reorientation was outside the coverage bounds of the optimization procedure. In cases with such severe deformity, the surgeon may not be able to correct the acetabulum to a radiographically normal orientation, and the optimization procedure may need to be adjusted to enforce a maximum achievable correction based on the original degree of deformity.

While this computational approach was able to identify acetabular orientations that optimize joint mechanics, it is currently unable to account for two important clinical concerns. First, the optimization algorithm is unable to detect whether a given acetabular orientation is likely to cause femoroacetabular impingement, which has been shown to adversely affect PAO survivorship [3, 40]. Numerous commercial and research software algorithms have been developed that predict impingement-free range-of-motion for planning PAO corrections [4, 22] or alleviating femoroacetabular impingement [29, 36], and such software is becoming increasingly utilized by clinicians when planning hip preservation procedures to ensure that improved acetabular coverage does not create secondary bony impingement that would limit a patient’s range-of-motion. Incorporating a similar collision detection technique should be a primary focus of future studies using this optimization technique. Similarly, the optimization procedure is currently unable to account for concurrent femoral procedures. The preoperative femur surface was used for all patient models in this work, and the optimal acetabular orientations were identified based on the articulation between the reoriented acetabular fragment and the preoperative femoral geometry. However, recent computational models suggest that if left unaddressed, femoral head-neck offset deformity may adversely affect joint contact stress after PAO [32]. Combined with clinical experience, such findings have led many hip preservation surgeons to address both sides of the joint concurrently through addition of arthrotomy or arthroscopy with femoral osteochondroplasty. Out of 107 PAOs performed at our institution between January 2018 and December 2020 for treatment of hip dysplasia, 84 (78.5%) had concurrent hip arthroscopy. It is possible that if the postoperative femoral geometry had been used for the patients that underwent concurrent hip arthroscopy, different optimal reorientations may have been identified. Future work will need to investigate the effects of concurrent femoral procedures on the optimal acetabular orientations identified using this optimization technique.

This work has several additional limitations that warrant further discussion. First, the DEA methodology used in this work makes several simplifying assumptions in relation to rigid bone, exclusion of the acetabular labrum, and cartilage material properties [37, 38]. Furthermore, while the loading parameters applied to the models in this study were obtained from patients with hip dysplasia [15], previous work has demonstrated sensitivity of DEA-computed contact stresses to applied loading [37], indicating that applying an average loading scheme to all patient models may not be accurate depending on patient-specific joint deformity and gait compensation mechanisms. However, given that motion capture data was not available for these patients, the study is limited to application of average gait information. While we elected to load our DEA models with average walking gait parameters, it is possible that loading our models with movements other than walking gait, such as a sit-to-stand task or deep squatting maneuvers, would result in the prediction of a different acetabular orientation as being mechanically or clinically optimal. Future studies should consider the effects of additional loading schemes on the predicted optimal acetabular corrections. Finally, the chronic contact stress-time exposure metric used in this work was scaled by patient age, which implies that hip joint loading has not changed since birth. This is likely inaccurate, particularly for the timespan prior to skeletal maturity, but provided the best approximation for chronic loading given that patient age at the time of surgery was the only information available.

Overall, this computational approach found unique, patient-specific acetabular orientations were needed to optimize joint contact mechanics. Optimizing based solely on minimizing chronic exposures frequently predicted optimal acetabular corrections that would lead to femoroacetabular impingement during normal, physiologic hip range of motion. Implementing a surgical rotation term in the cost function produced optimal orientations that were more realistic from a clinical perspective. However, even the clinically optimal orientations trended toward what would be considered acetabular over-coverage, and performing such corrections may pose a risk for secondary femoroacetabular impingement. The increased lateral and anterior coverage in the clinically optimal corrections compared to the surgically achieved corrections may explain the significantly improved contact mechanics of the optimized reorientations compared to those achieved surgically. However, greater radiographic coverage did not always guarantee a greater improvement in contact mechanics, and some clinically optimal reorientations were found with similar coverage to what was achieved surgically. These results indicate that achieving an optimal patient-specific correction, rather than just a radiographically normal correction, may be necessary to improve contact mechanics in dysplastic hips and reduce the risk of OA progression after PAO.

## Data Availability

All data produced in the present study are available upon reasonable request to the authors.

## ACKNOWLEDGMENTS

We thank the Orthopaedic Research and Education Foundation and the National Institute of General Medical Sciences of the National Institutes of Health for supporting this work under respective award numbers Career Development Grant 17-001 and R25GM058939.

## APPENDIX

### Mechanically Optimal Exposure

The mechanically optimal exposure metric was defined as

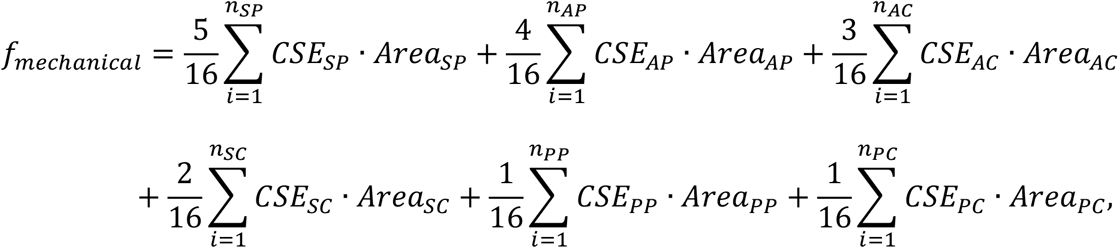

where *CSE* is the contact stress over-exposure magnitude, in MPa-years, at individual triangular facet *i* on the DEA model surface, and *Area* is the contacting surface area, in mm^2^, of facet *i*. Subscripts refer to individual subregions of the acetabulum (*SP* = superolateral peripheral; *AP* = anterior peripheral; *PP* = posterior peripheral; *SC* = superolateral central; *AC* = anterior central; *PC* = posterior central).

### Clinically Optimal Exposure

The clinically optimal exposure metric was defined as

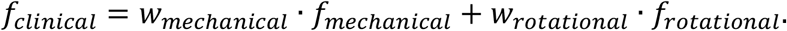

In this calculation, *f*_*mechanical*_ is the original, mechanically optimal exposure cost function, and *f*_*rotational*_ is a component associated with the surgical rotation of the acetabular fragment:

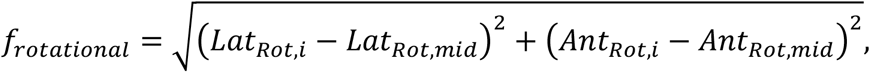

where *Lat*_*Rot*,*i*_ is the lateral acetabular coverage, in degrees, at the *i*th candidate reorientation; *Lat*_*Rot*,*mid*_ is the mid-range lateral acetabular coverage (i.e., 34 degrees); *Ant*_*Rot*,*i*_ is the anterior acetabular coverage, in degrees, at the *i*th candidate reorientation; and *Ant*_*Rot*,*mid*_ is the mid-range anterior acetabular coverage (i.e., 32 degrees). Given the very different measurement scales of *f*_*mechanical*_ (in MPa-years-mm^2^) and *f*_*rotational*_ (in degrees), weights *w*_*mechanical*_ and *w*_*rotational*_ were implemented to balance their relative contributions, and these two weights were determined on a patient-specific basis by iteratively adjusting both until an optimal reorientation was identified that did not fall along the limits of acetabular coverage (i.e., *f*_*mechanical*_ was dominant) or at the exact coverage midpoint (i.e., *f*_*rotational*_ was dominant).

